# Lifelong impacts of puberty timing on human plasma metabolic profiles

**DOI:** 10.1101/2023.09.09.23295295

**Authors:** Zengjun Li, Si Fang, Dong Liu, Fei Li, Cairong Zhu, Jian Zhao

**Affiliations:** Department of Epidemiology and Biostatistics, West China School of Public Health and West China Fourth Hospital, Sichuan University, 610041, China; Population Health Sciences, Bristol Medical School, University of Bristol, Bristol, United Kingdom; Medical Research Council (MRC) Integrative Epidemiology Unit (IEU), University of Bristol, Bristol, United Kingdom; Ministry of Education and Shanghai Key Laboratory of Children’s Environmental Health, Xinhua Hospital, Shanghai Jiao Tong University School of Medicine, Shanghai, 200092, China; Department of Developmental and Behavioral Pediatric & Child Primary Care, Brain and Behavioral Research Unit of Shanghai Institute for Pediatric Research, Xinhua Hospital, Shanghai Jiao Tong University School of Medicine, Shanghai, China; Department of Maternal and Child Health, School of Public Health, Shanghai Jiao Tong University, 200092, China

## Abstract

There has been uncertainty regarding the long-term impact of puberty timing on human plasma metabolites. This lack of clarity can be attributed to the influence of confounding factors present in conventional observational studies. To determine the causal effect of puberty timing on plasma metabolites, we employed a two-sample Mendelian randomization (MR) analysis, complemented by MR mediation analysis assessing the direct effect. We utilized data from a large-scale meta-analysis of genome-wide association studies (GWAS) on puberty timing, consisting of 329,345 women of European ancestry, and a meta-analysis of GWAS on plasma metabolites, involving up to 86,507 individuals. Our findings provide moderate evidence supporting a causal effect of puberty timing on 23 out of 174 plasma metabolites. After excluding 7 single nucleotide polymorphisms (SNPs) related to birth weight and childhood adiposity, causal effects remained for 16 metabolites. Through two-step MR analysis, we observed strong evidence that adulthood adiposity mediated the causal relationships of puberty timing on 35 plasma metabolites. We also observed moderate evidence for an independent causal effect of puberty timing on 10 metabolites through multivariable MR analysis. We further used metabolomic data measured in the UK Biobank (UKB) to perform a replication analysis to validate the causal effect estimated. Nine amino acids were identified in the UKB, and the replication analysis supported our main findings.

## Introduction

The onset of puberty, influenced by a combination of genetic and environmental factors ^1,2^, plays a pivotal role in shaping growth and development throughout an individual’s lifespan ^3^. Early puberty onset has been associated with various unfavorable health outcomes, including childhood and adulthood obesity ^4,5^, type 2 diabetes ^6^, hypertension ^7^, breast cancer ^8^, and mental disorders ^9^. The hormonal fluctuations during puberty initiation trigger metabolic changes that can have significant implications ^3^. The investigation of the causal relationship between puberty timing and plasma metabolites in adulthood is of utmost importance, given the increasing recognition of plasma metabolites as effective biomarkers for illness diagnosis and prognosis ^10,11^. This exploration will provide insights into the long-lasting effects of early-life determinants, such as puberty onset, on human metabolism later in life. Unfortunately, previous studies on this subject have been limited, predominantly relying on conventional multivariable analysis methods that are susceptible to confounding or reverse causation biases ^5,12,13^. Additionally, the utilization of longitudinal studies for the examination of the correlation between puberty timing and the concentration of plasma metabolites necessitates an extended duration of follow-up, resulting in limited empirical support.

Mendelian randomization (MR) is an increasingly used study design that employs genetic variants as instrumental variables to draw causal inferences between an exposure of interest (e.g., puberty timing) and an outcome (e.g., a metabolite) ^14^. MR analyses are much less vulnerable to conventional confounding as a result of the random assignment of alleles during meiosis ^15^, hence differentiating MR from normal observational research. Moreover, MR analyses are less prone to reverse causation because the assortment of alleles precedes the initiation of exposure (e.g., puberty timing).

A recent MR study investigated if puberty timing has a distinct influence on adulthood cardiometabolic traits ^16^. The results suggested that the effects of puberty timing on adulthood adiposity and cardiometabolic traits are not likely driven by itself but by childhood adiposity. Additionally, it is worth noting that the majority of the cardiometabolic characteristics examined in this study mostly focused on lipids, resulting in a very restricted range of metabolites investigated. Consequently, in this study, we aimed to examine whether puberty timing is likely to, directly and/or indirectly, impact a wide range of plasma metabolites, including amino acids, biogenic amines, acylcarnitines, phosphatidylcholines, lysophosphatidylcholines, sphingomyelins, and hexose in adulthood in a two-sample MR framework. Along with conducting univariable MR analysis to infer the total effect of puberty timing on plasma metabolites, we also performed MR mediation analysis to take the mediating effect of adulthood adiposity into account.

## Results

### Lifelong causal effects of puberty timing on human metabolites

The study design is illustrated in **Fig 1**. In the primary analysis, all genetically significant SNPs were used as instrument variables. Additionally, to minimize the bias arising from horizontal pleiotropy, a secondary analysis was also performed by excluding SNPs that were genetically associated with confounders (e.g., birth weight and childhood adiposity) (**Fig 2**). Two-sample MR analysis suggested moderate causal associations between genetically predicted puberty timing and 23 plasma metabolites within the subclasses including acylcarnitines, amino acids, biogenic amines, and lysophosphatidylcholines based on the primary analysis (i.e., inverse-variance weighted (IVW) analysis) results (**Fig 3-4; Table S7**) with *P* values ranging from 9.09 × 10^-4^ to 4.92 × 10^-2^. The MR analysis results supported positive causal associations of later puberty timing with higher levels of 20 metabolites. The strongest causal evidence was found for dodecanedioylcarnitine (beta = 0.11, 95% CI: 0.04 to 0.17, *P* = 9.09 × 10^-4^), which means each year delayed puberty onset was associated with 0.11 standard deviation (SD) units higher of dodecanedioylcarnitine. By contrast, lower levels of three amino acids, aspartate (beta = −0.04, 95% CI: -7.19 × 10^-2^ to - 6.40 × 10^-4^, *P* = 4.60 × 10^-2^), phenylalanine (beta = −0.03, 95% CI: -5.03 × 10^-2^ to - 1.20 × 10^-3^, *P* = 3.98 × 10^-2^), and tyrosine (beta = −0.03, 95% CI: -5.17 × 10^-2^ to -8.46 × 10^-3^, *P* = 6.41 × 10^-3^), were observed per one year later onset of puberty.

**Fig 1.**
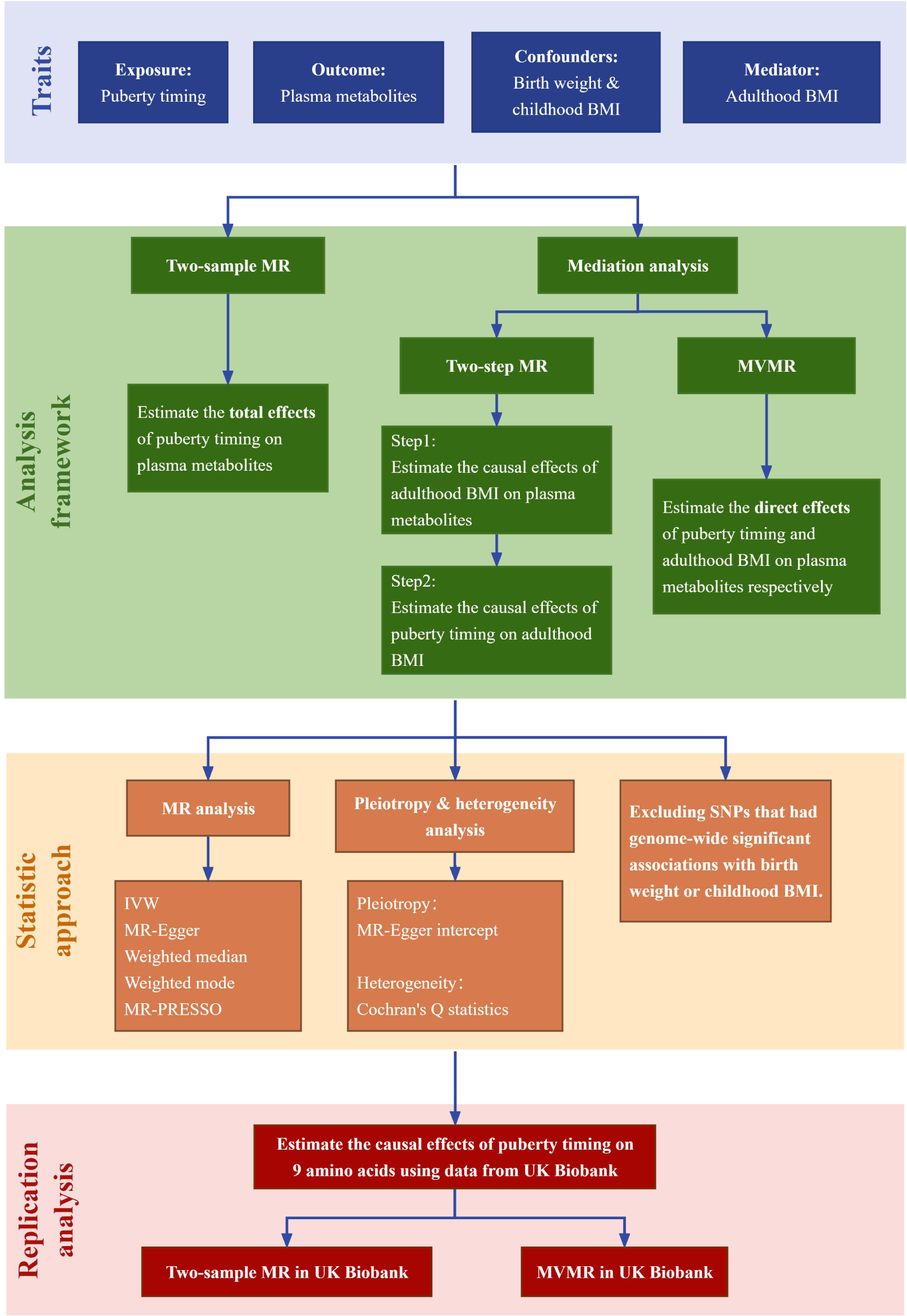
The schematic diagram for Mendelian randomization of puberty timing on plasma metabolites. The primary analysis was conducted utilizing a two-sample MR (Mendelian randomization) approach to investigate the total effect of puberty timing on plasma metabolites. In addition, we employed the two-step MR and MVMR (multivariable Mendelian randomization) approach, taking the mediating effect of adulthood BMI (body mass index) into account to infer the “direct” and “indirect” effects. The IVW (inverse-variance weighted) approach was used as the primary analysis, while MR-Egger, weighted median, weighted mode, and MR-PRESSO (Mendelian randomization pleiotropy residual sum and outlier) were used as sensitivity analyses. Subsequently, we excluded SNPs (single nucleotide polymorphisms) that were genetically associated with birth weight and/or childhood BMI from genetic instruments to minimize the risk of bias due to horizontal pleiotropy. We further used metabolomic data obtained from the UK Biobank to perform a replication analysis, aiming to validate the causal effect estimated.

**Fig 2.**
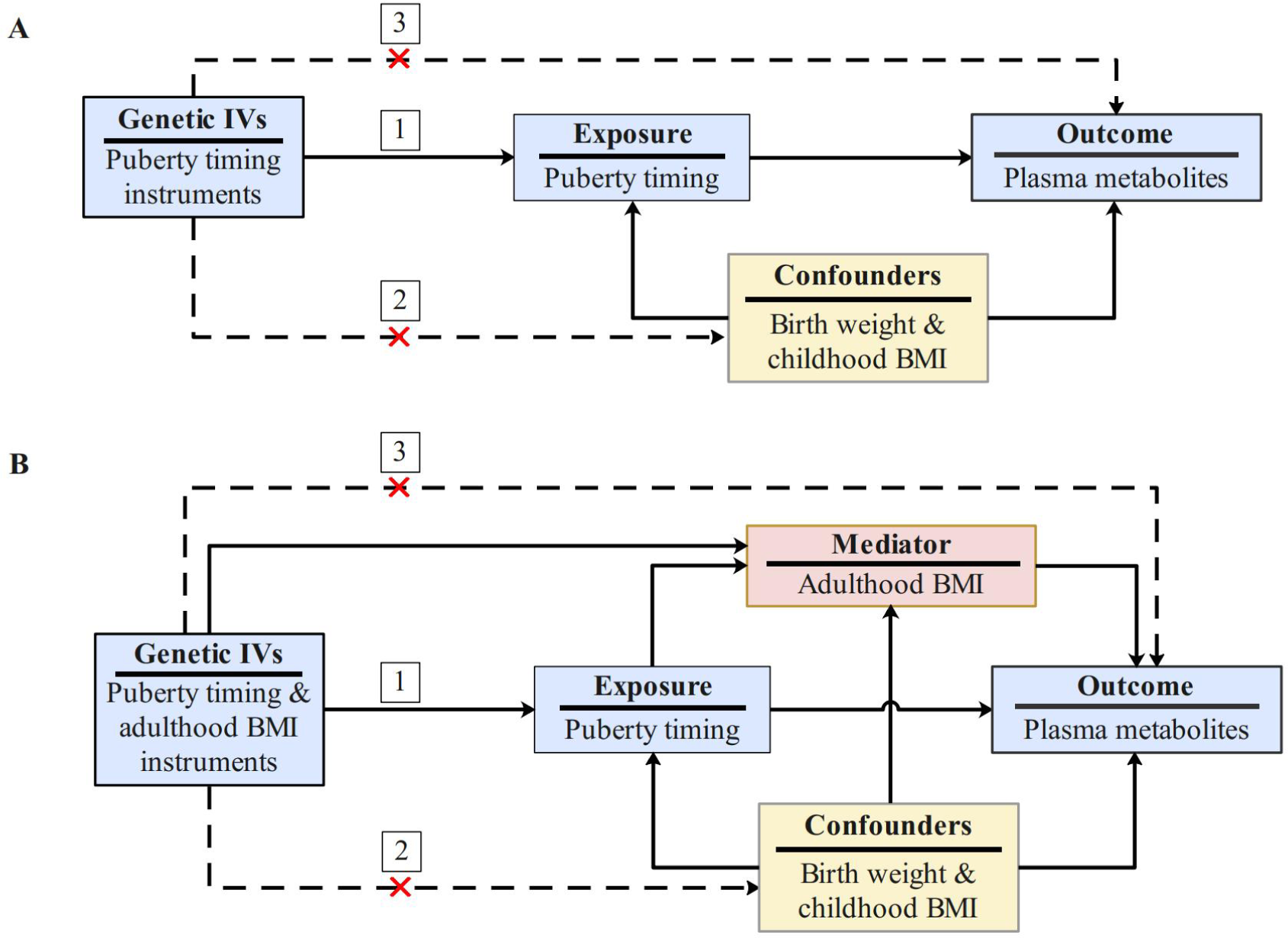
The directed acyclic graphs of the Mendelian randomization study. The figures depict directed acyclic graphs for univariable Mendelian randomization (MR) and mediation MR analysis in panel A and panel B respectively. The dashed lines represent potential horizontal pleiotropy or direct causal effects between variables, which would violate the MR assumption. MR approach is based on three assumptions: (1) the IVs (instrument variables) are strongly associated with the exposure (e.g., puberty timing); (2) the genetic IVs are not associated with any confounders (e.g., birth weight and childhood BMI (body mass index)); (3) the genetic IVs should influence the outcome (e.g., a metabolite) through the exposure independently instead of through other pathways. The primary analysis was performed, wherein SNPs that exhibited genetic associations with puberty timing were selected as IVs. To minimize the risk of bias due to horizontal pleiotropy, a secondary analysis was conducted wherein SNPs that exhibited genetic associations with birth weight and/or childhood BMI were excluded.

**Fig 3.**
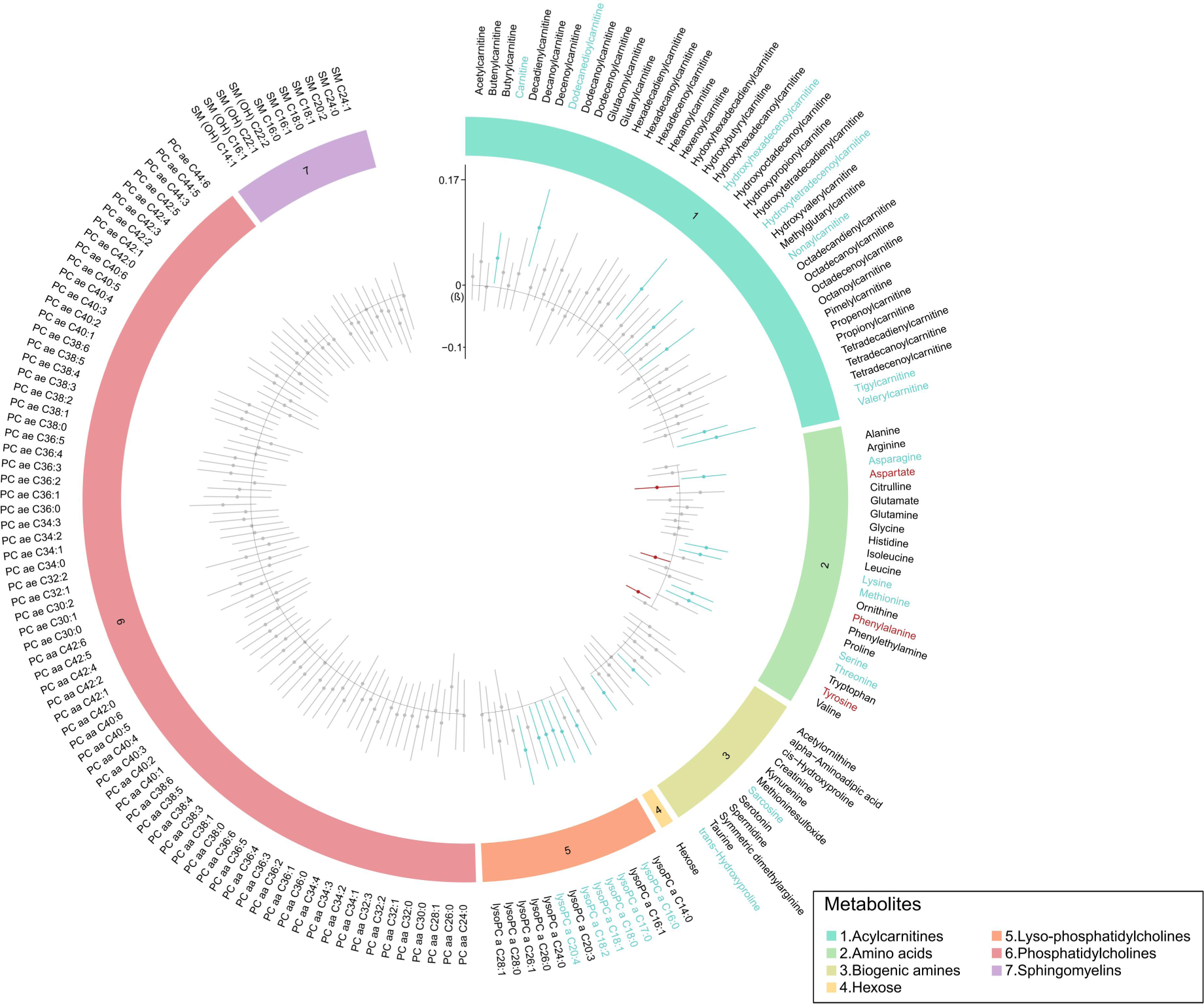
Circos plot depicting the causal effect estimates of puberty timing on all 174 plasma metabolites. The legend located at the lower section of the figure provides a visual representation of distinct subclasses of plasma metabolites. The presence of metabolites highlighted with the color blue indicates a positive causal effect of puberty timing on the concentration of plasma metabolites. Specifically, each year later in puberty timing is associated with higher levels of metabolites. In contrast, the identification of metabolites highlighted with the color red suggests a negative causal effect of puberty timing on the concentration of plasma metabolites, that is each year later in puberty timing is associated with lower levels of metabolites. The presence of metabolites that had been designated as black indicates that there is insufficient evidence to establish a causal relationship. The evidence indicates a causal effect of puberty timing on 23 plasma metabolites. Error bars represent 95% confidence intervals. All statistical tests were two-sided.

**Fig 4.**
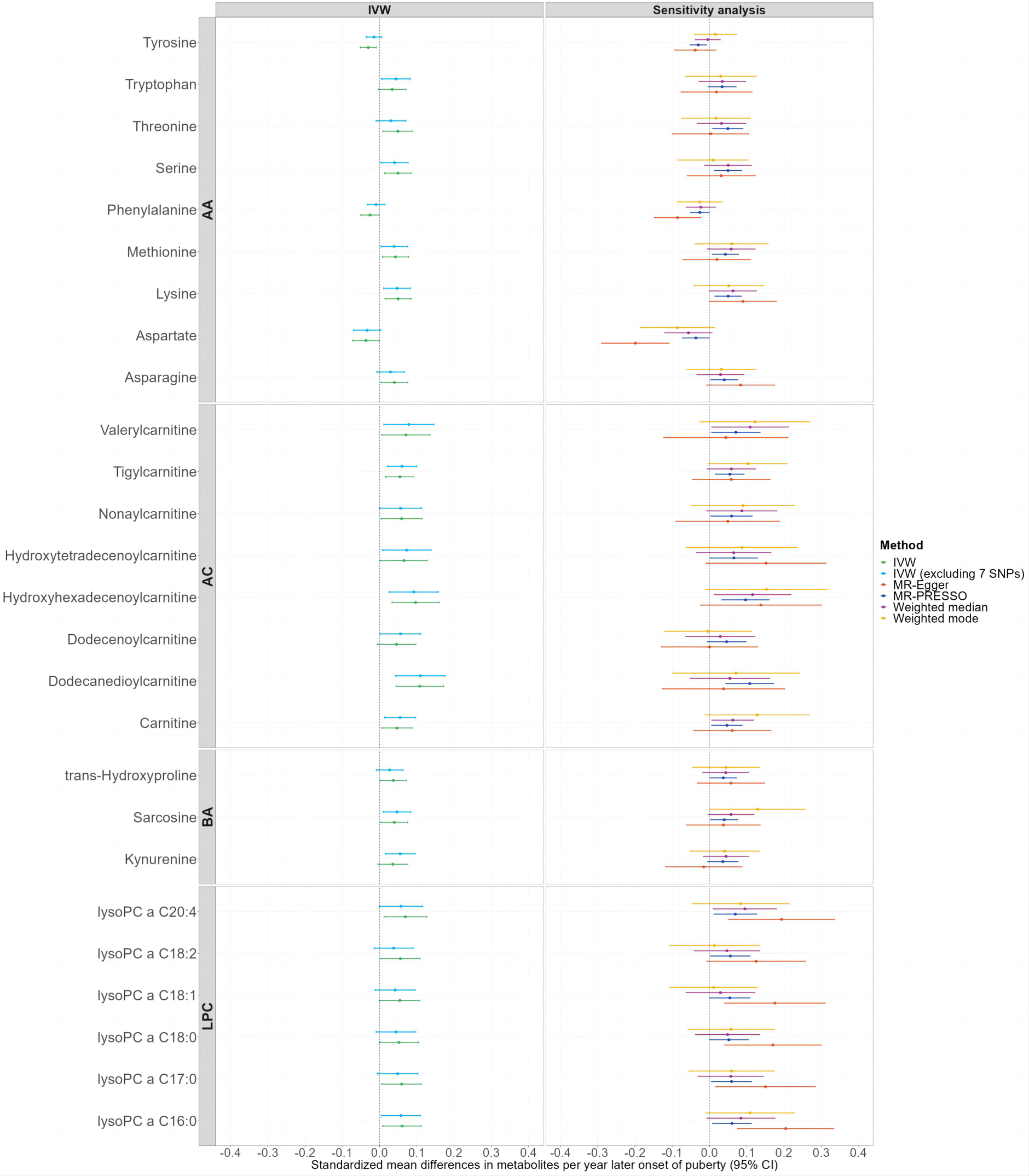
Forest plots showing the causal effect estimates of puberty timing on plasma metabolites. The legend located on the right side of the forest plot indicates different analysis approaches. Plasma metabolites reaching nominal significance (P < 0.05) were shown according to the results of IVW (inverse-variance weighted). The results presented on the left side of the plot were analyzed using the IVW approach. The colors represent green, using the IVW method to estimate the effect size of puberty timing on plasma metabolites; blue; using the IVW method, but excluding 7 genetic instruments genetically related to birth weight and/or childhood BMI (body mass index). The results presented on the right were derived from four distinct sensitivity analyses. The sensitivity analysis findings indicate that the results of the primary analysis are largely robust. Error bars represent 95% confidence intervals. All statistical tests were two-sided. The metabolites were divided according to different subclasses. AA amino acids; AC acylcarnitines; BA biogenic amines; LPC lysophosphatidylcholines.

For the 23 metabolites indicating moderate causal consequences of puberty timing, causal effect estimates in sensitivity analyses were broadly consistent with the results of the primary IVW analysis (**Table S8-S9**). Of note, intercept terms estimated in the MR-Egger regression analysis suggested that some of the results might be biased by horizontal pleiotropy including aspartate, phenylalanine, alpha-aminoadipic acid, lysoPC a C16:0, PC aa C38:0 and PC aa C40:1, which was also supported by evidence of heterogeneity tested via Cochran’s Q statistic in the IVW analysis (**Table S10-S11**).

After excluding 7 SNPs associated with birth weight and childhood body mass index (BMI), the secondary IVW analysis confirmed moderate evidence for causal associations of later puberty timing with higher levels of 16 metabolites out of the aforementioned 23 metabolites in the primary two-sample MR analysis. It is noteworthy that 3 additional metabolites, including dodecenoylcarnitine: beta = 0.06, 95% CI: 2.75 × 10^-3^ to 1.10 × 10^-2^, *P* = 3.94 × 10^-2^; tryptophan: beta = 0.04, 95% CI: 5.61 × 10^-3^ to 8.25 × 10^-2^, *P* = 2.47 × 10^-2^; and kynurenine: beta = 0.06, 95% CI: 1.57 × 10^-2^ to 9.58 × 10^-2^, *P* = 6.38 × 10^-3^, appeared to be impacted by puberty timing in the secondary IVW analysis, after controlling for potentially horizontal pleiotropic effects via birth weight or childhood adiposity. On the contrary, 10 out of the 23 putative metabolites observed in the primary analysis turned to statistically non-significant in the secondary IVW analysis (**Fig 4, Table S12**), which included phenylalanine, threonine, asparagine, aspartate, tyrosine, trans-hydroxyproline, lysoPC a C17:0, lysoPC a C18:0, lysoPC a C18:1, and lysoPC a C18:2. Among them, intercept terms estimated in the MR-Egger regression analysis suggested possible horizontal pleiotropy for phenylalanine and heterogeneity for aspartate and lysoPC a C18:1 (**Table S13-S14**).

## Mediation analysis

The estimated causal effects of adulthood BMI on 174 metabolites suggested that adulthood BMI strongly influenced plasma concentrations of some metabolites through two-step MR analysis, including 35 metabolites reaching nominal significance and 13 reaching statistical significance after the multiple testing Bonferroni correction (**Table S15**). Further, we found a causal effect of puberty timing (beta = −0.13, 95% CI: -0.17 to -0.09, *P* = 8.71 × 10^-12^) on the mediator (i.e., adulthood BMI). By exploiting the “product of coefficients” method, we derived the indirect effects of puberty timing on each plasma metabolite (**Table S15**). For instance, we found that each year of delayed puberty onset was associated with 0.01 SD units increase in serine through the pathway of adulthood adiposity (beta = 0.01, 95% CI: 1.04 × 10^-3^ to 2.44 × 10^-2^, *P* = 0.03), indicating that adulthood adiposity was a putative factor that mediated the causal relationships of puberty timing on certain metabolites. To further rule out the influence of horizontal pleiotropy, we removed a total of 13 SNPs highly related to childhood adiposity and birth weight from adulthood BMI and puberty timing genetic instrument sets. A total of 31 metabolites reached nominal significance and 8 reached the multiple testing Bonferroni corrected significance level. Moreover, the effect size of puberty timing on adulthood adiposity attenuated from -0.13 kg/m^2^ to -0.09 kg/m^2^ (beta= −0.09, 95% CI: -0.11 to -0.06, *P* = 1.81 × 10^-8^). Nevertheless, the mediating role of adulthood BMI between puberty timing and adulthood metabolites persisted when the potential pleiotropic effects were adjusted for (**Table S17**).

The MVMR analysis results supported causal evidence for direct causal effects of puberty timing on 10 metabolites (i.e., carnitine, dodecanedioylcarnitine, hydroxytetradecenoylcarnitine, tigylcarnitine, lysine, ornithine, cis-hydroxyproline, trans-hydroxyproline, lysoPC a C20:4, and PC aa C40:4) based on nominal significance threshold of *P* < 0.05. Moreover, among the 10 significant metabolites, the results suggested that carnitine, dodecanedioylcarnitine, tigylcarnitine (acylcarnitines), lysine (amino acids), and lysoPC a C20:4 (lysophosphatidylcholines) were directly affected by puberty timing and had the same direction as results in the primary analysis (**Fig 5; Table S16**). Notably, the effect sizes attenuated to varying degrees, because adulthood BMI was adjusted for in the MVMR analysis. For example, the coefficient for dodecanedioylcarnitine attenuated from 0.11 SD units to 0.07 SD units per year later onset of puberty, approximately half of the original estimate. Compared with the results in the primary MR analysis, except for phenylalanine and tyrosine (amino acids) which showed inconsistent directions, the remaining 21 metabolites showed the same direction as the primary two-sample MR analysis. Furthermore, the MVMR analysis suggested that the plasma levels of 17 metabolites (i.e., hydroxyhexadecenoylcarnitine, nonaylcarnitine, valerylcarnitine, asparagine, aspartate, methionine, phenylalanine, serine, threonine, tyrosine, sarcosine, trans-hydroxyproline, lysoPC a C16:0, lysoPC a C17:0, lysoPC a C18:0, lysoPC a C18:1, lysoPC a C18:2) were mediated by adulthood BMI rather than directly affected by puberty timing (**Table S16; Table S18**).

**Fig 5.**
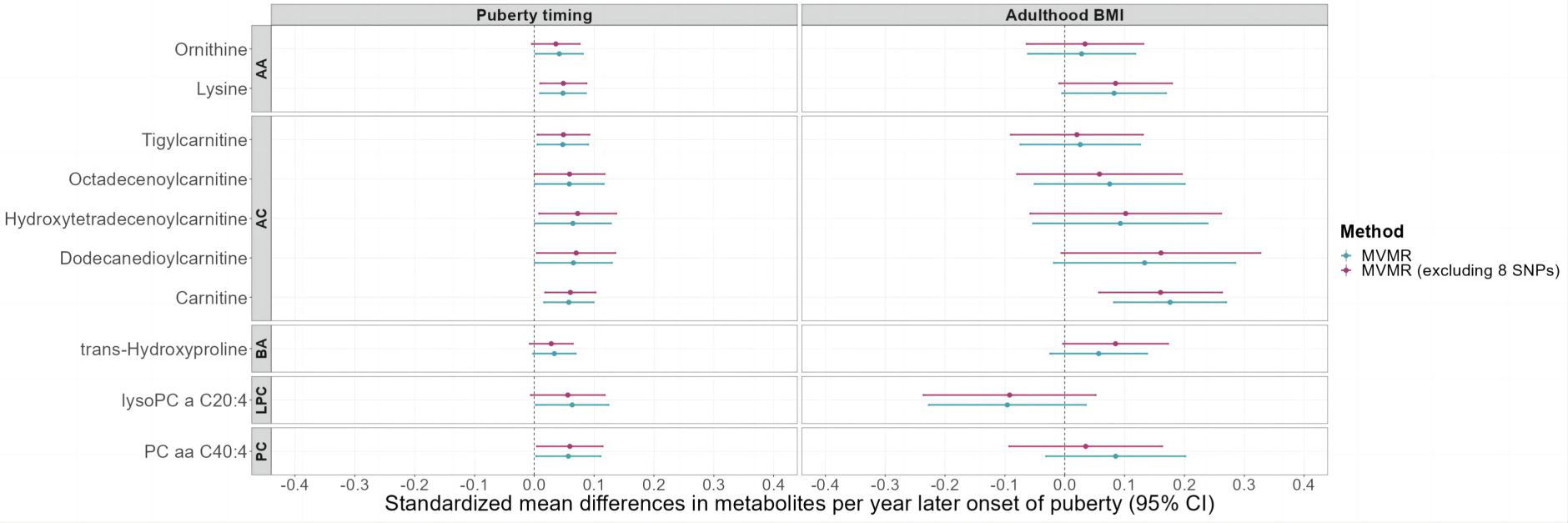
Forest plots comparing the effect estimates of puberty timing and adulthood BMI on plasma metabolites in MVMR analyses. The legend located on the right side of the forest indicates the distinct analysis. Plasma metabolites reaching nominal significance (*P* < 0.05) in puberty timing were shown according to the results of the MVMR (multivariable Mendelian randomization) approach. The colors represent blue, using the MVMR method to estimate the effect size of puberty timing or adulthood BMI on plasma metabolites; purple; using the MVMR method, but excluding 8 genetic instruments associated with birth weight and/or childhood BMI (body mass index). The results on the left show the effect size of puberty timing on each plasma metabolite, whereas the results on the right present the effect size of adulthood BMI on each plasma metabolite. Error bars represent 95% confidence intervals. All statistical tests were two-sided. The metabolites were also divided according to different subclasses. AA amino acids; AC acylcarnitines; BA biogenic amines; LPC lysophosphatidylcholines; PC phosphatidylcholines.

## Replication analysis

We attempted to replicate the effect estimates for puberty timing on the identified nine metabolites in the UKB. In the two-sample MR analysis, a nominally significant causal relationship was revealed between puberty timing and two amino acids, namely phenylalanine (beta = −0.03, 95% CI: -5.38 × 10^-2^ to -2.45 × 10^-3^, *P* = 3.18 × 10^-2^) and valine (beta = −0.04, 95% CI: -6.54 × 10^-2^ to -3.82 × 10^-3^, *P* = 2.77 × 10^-2^, although none of the nine amino acids reached the strictly adjusted *P* threshold of 5.56 × 10^-3^ (0.05/9) (**Fig 6; Table S19**). Causal effect estimates in the replication analysis of the UKB data were consistent with those estimated in the primary two-sample MR analysis except for isoleucine and glutamine. Notably, consistent with the primary MVMR analysis results, in the MVMR analysis using the UKB data, the causal effects attenuated towards null after adjusting for adulthood adiposity, suggesting little direct effects were observed between puberty timing and the nine amino acids (**Fig 6; Table S20**).

**Fig 6.**
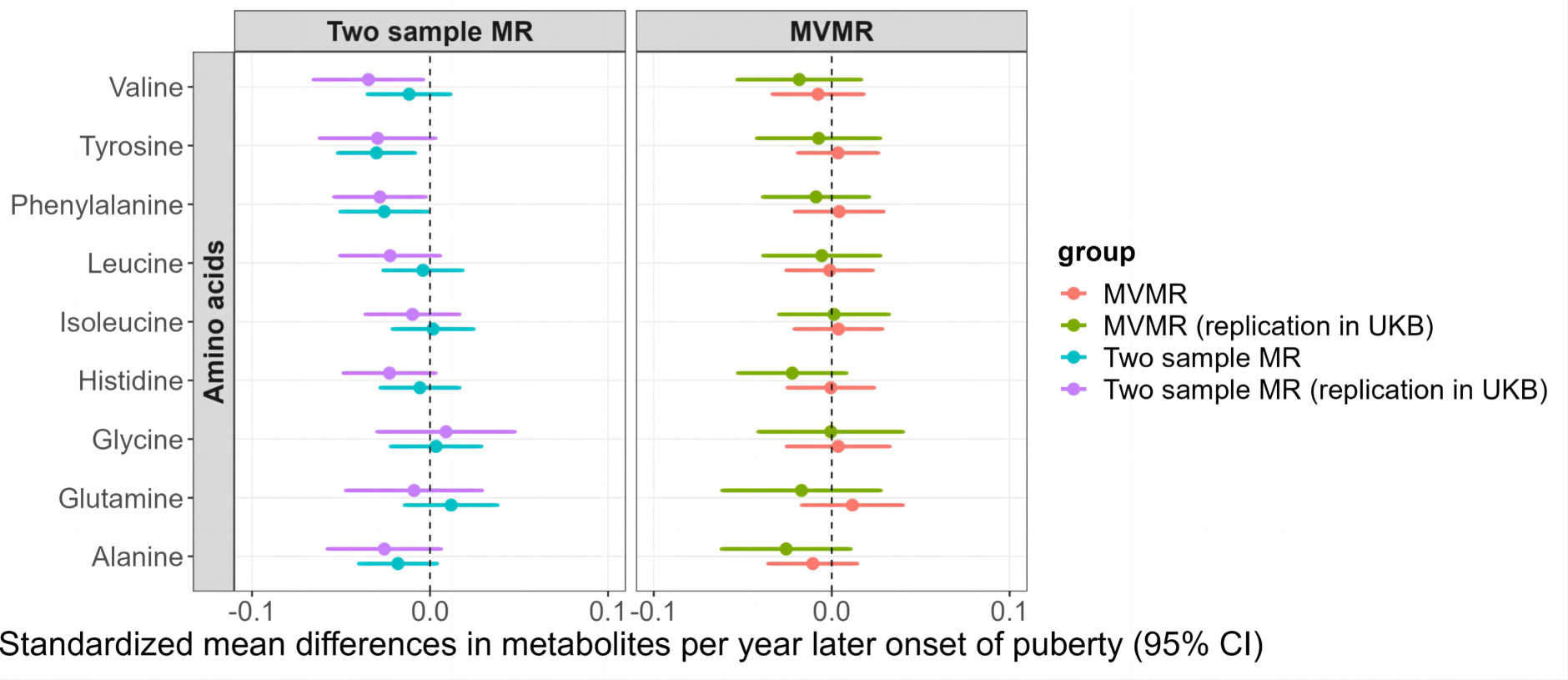
Forest plots comparing the effect estimates of puberty timing on nine amino acids in replication analyses. The legend located on the right side of the forest plot indicates different analysis methods. The results of two-sample MR (Mendelian randomization) and MVMR (multivariable Mendelian randomization) represented the total (blue) and direct impact (orange) of puberty timing on plasma metabolites, respectively. To inspect the robustness of our two-sample MR and MVMR analysis results (purple and green), we estimated the causal relationship of puberty timing on nine identified amino acids using data from the UKB (UK Biobank). Our findings in the primary MR analysis were further supported by the findings from external replication analysis in the UKB. Error bars represent 95% confidence intervals. All statistical tests were two-sided.

## Discussion

Plasma levels of metabolites have been investigated as promising biomarkers for disease risk stratification and critical molecular targets for developing novel treatments and interventions for a range of diseases and conditions ^17-20^. Given that puberty timing is consistently associated with the risk of developing various chronic diseases in adulthood ^21^, further causal insights into the relationship between puberty timing and plasma levels of metabolites will help understand underlying biological mechanisms and identify possible putative target biomarkers for disease treatment or intervention. Here, we systematically and extensively investigated the lifelong impacts of puberty timing on human plasma metabolic profiles, by integrating data on human puberty timing and plasma metabolism in the MR causal inference framework. Genetically predicted puberty timing appeared to have causal effects on 23 plasma metabolites in adulthood. After considering the potential mediating role of adulthood adiposity, direct causal effects of puberty timing on 10 adulthood metabolites remained, suggesting the causal effects of puberty timing on certain metabolites in adulthood were mediated by adulthood adiposity. Our findings in the primary MR analysis were further supported by the results from the mediation analysis and external replication analysis in the UKB.

Among the 23 plasma metabolites identified in the two-sample MR analysis, seven belong to acylcarnitines. In MVMR analysis, 5 of the 10 metabolites affected by puberty timing were also confirmed within the subclass of acylcarnitines, including carnitine, tigylcarnitine (short-chain), dodecanedioylcarnitine, hydroxytetradecenoylcarnitine, and octadecenoylcarnitine (medium- and long-chain). With the causal effect estimates ranging from 0.05 SD to 0.07 SD per year of delayed puberty onset, our findings suggested that certain acylcarnitine metabolites were directly and independently impacted by puberty timing. Acylcarnitines were a group of compounds formed from carnitine and acyl-CoAs in mitochondria or peroxisomal by carnitine acyltransferases ^22,23^, which can cross mitochondrial and cell membranes. Short-chain acylcarnitines are generated from the degradation of glucose, amino acids, and fatty acids. Medium- and long-chain acylcarnitines are largely produced from fatty acids participating in the transportation of fatty acids for the following fatty acids oxidation ^24^. It has been well understood that increased plasma levels of acylcarnitine are associated with a higher risk of cardiovascular disease ^25-27^. The underlying mechanism might be the accumulation of acylcarnitine was associated with a disturbed β-oxidation rate and mitochondrial dysfunction ^28,29^. In addition, previous studies have shown that a shorter reproductive lifespan is associated with an increased risk of cardiovascular disease events ^30^. Furthermore, in our study, we found a positive causal effect of delayed puberty timing on the accumulation of acylcarnitine. Taking together, our findings may shed new light on the correlation between puberty timing and cardiovascular diseases. However, whether puberty timing affects cardiovascular diseases through the acylcarnitine metabolism pathway has not been illuminated, thus more studies are warranted.

Of note, we found 8 amino acids in adulthood were significantly impacted by puberty timing in the primary MR analysis, among which inverse causal effects of puberty timing on the concentrations of tyrosine and phenylalanine were externally validated using data from the UKB. After accounting for adulthood adiposity in the MVMR analysis, the direct causal effects of puberty timing on these 8 amino acids attenuated to null in both the primary and replication MVMR analysis. It suggests a mediating role of adulthood adiposity in the causal pathway from puberty timing to the aforementioned amino acids. Our findings were partly consistent with the findings from a previous study ^16^, where an inverse association between age at menarche and phenylalanine measured at 18 years old was revealed in the conventional observational analysis among the females in the Avon Longitudinal Study of Parents and Children (ALSPAC), but a causal association was failed to be established in the one-sample MR analysis using the same ALSPAC cohort data.

In our two-step MR analysis, we found that adulthood BMI influenced the levels of 35 metabolites, and adulthood BMI itself was also influenced by puberty timing. After adjusting for adulthood BMI through MVMR analysis, the number of metabolites influenced by puberty timing decreased from 23 to 10. Our findings suggested that adulthood BMI may serve as a mediator between puberty timing and plasma metabolites. The reason lies in the fact that early-maturing individuals may face a higher risk of obesity and exhibit a higher likelihood of metabolic abnormalities in adulthood ^31^. Obesity can also significantly increase the risk of developing metabolic disorders by affecting energy metabolism and altering pathways ^32^, which could be the reason for changes in acylcarnitine and amino acid metabolism.

The strengths of this study were as follows. First, the data we used were curated from the latest and largest GWAS in European ancestry. Second, the MR mediation analysis makes it possible to infer the direct and indirect effects of genetically predicted puberty timing on adulthood plasma levels of metabolites. In the two-step MR analysis, we used adulthood BMI as a mediator to investigate the indirect effects of puberty timing on metabolites. As for MVMR analysis, we included both adulthood BMI and puberty timing as exposures to explore the direct effect of puberty timing on metabolites. Third, to minimize the risk of bias due to horizontal pleiotropy, we excluded SNPs that had genome-wide significant associations with birth weight or childhood obesity. Finally, external replication analysis using data from the UKB supported our primary analysis results, suggesting the robustness of our findings.

Limitations were also noted. First, our study results were primarily based on GWAS conducted in individuals of European descent. Consequently, it should be cautious when generalizing the results to non-European populations. Second, despite extensive analyses were performed to minimize the risk of bias due to horizontal pleiotropy, it cannot be fully ruled out, thus the results should be interpreted with caution. Third, the puberty timing GWAS were conducted in females exclusively, while the metabolites GWAS included female and male individuals. As there may be sex- specific effects on the levels of metabolites ^33^, this could potentially impact our causal effect estimates. Thus, future studies taking into account the sex-specific effects with regard to the relationship between puberty timing and human plasma metabolism are warranted.

In summary, we explored the causal relationship of puberty timing with human plasma metabolite levels using publicly available GWAS summary data. We found a causal effect of puberty timing on 23 plasma metabolites through two-sample MR analysis, and most of the effects were mediated by adulthood adiposity. The findings provide novel insights into the causal inferences of puberty timing on human metabolism. Additionally, the role of adulthood adiposity in mediating the causal relationship between puberty timing and plasma metabolites is identified.

## Material and Methods

### Data sources

The information on all GWAS data used in the analyses is shown in **Table S21**. Summary statistics on age at menarche (AAM) (measured in years) were obtained from the largest meta-analysis of GWASs, including 329,345 women of European ancestry from 42 cohort studies ^34^. A total of 389 SNPs reaching genome-wide significance (*P* < 5 × 10^−8^) were identified. There exists a genetic correlation (r^g^=0.74) between the AAM of females and the age at voice breaking of males, which suggests that many genetic variants have comparable effects on puberty timing in both males and females. Consequently, there was credibility that genetic variants of AAM can provide confident insights into puberty timing in males as well ^35^. Thus, in our study, these genetic variants were considered instrumental variables for puberty timing. We also refer to AAM as puberty timing in the present study.

A total of 174 plasma metabolites in seven biochemical categories (i.e., amino acids, biogenic amines, acylcarnitines, phosphatidylcholines, lysophosphatidylcholines, sphingomyelins, and hexoses), from the largest GWAS of human plasma metabolites, were used as the outcomes in this study ^36^. Summary statistics on all metabolites measured by high-throughput platforms were obtained from the cross-platform meta-analysis GWAS of blood metabolites, which included several cohorts with sample sizes for each metabolite ranging from 8,569 to 86,507 individuals of European ancestry. The GWASs were undertaken in cohorts of Fenland study (N = 9,363), EPIC-Norfolk (European Prospective Investigation into Cancer-Norfolk) (N = 5,841), and INTERVAL study (Metabolon Discovery HD4 platform, N = 8,455 and ^1^H nuclear magnetic resonance (NMR), Nightingale, N = 40,905). The results were further meta-analyzed with two published studies conducted by Kettunen et al. (N = 24,925) and Shin et al. (N = 7,824) ^37,38^.

The adulthood adiposity GWAS conducted by the Genetic Investigation of Anthropomorphic Traits (GIANT) consortium included 315,347 adults of European ancestry, combining data from the Research Program on Genes, Environment, and Health (RPGEH), Genetic Epidemiology Research on Aging (GERA) cohort and a meta-analysis of GWASs conducted by Locke et al. ^39,40^. A total of 147 genome-wide significant variants associated with adulthood BMI were selected as genetic instruments in the MR mediation analysis.

Since birth weight and childhood adiposity have been reported to be associated with puberty timing and plasma metabolite level changes in previous studies ^41-43^, we excluded SNPs, which were genetically significantly associated with these two traits, from the genetic instruments for puberty timing to minimize the potential risk of bias due to horizontal pleiotropy. Full summary data on birth weight and childhood BMI were obtained from the Early Growth Genetics (EGG) Consortium ^43,44^. In the birth weight GWAS, Warrington et al. conducted a meta-analysis of the fetal genetic effects in up to 297,356 Europeans with their own birth weight and 210,248 Europeans with offspring birth weight ^43^. Bradfield et al. performed a trans-ancestral GWAS meta-analysis of childhood BMI from 30 studies, but only data on Europeans were used in the analyses ^44^.

We then attempted to replicate the findings of our study by using data from independent GWASs. Data on puberty timing were obtained from a GWAS conducted on up to 182,416 females of European descent from 58 studies ^45^. Summary statistics on adulthood BMI were obtained from a meta-analysis of 322,154 European ancestries from the GIANT consortium ^40^. Furthermore, we obtained summary statistics from a GWAS of metabolites measured in the UKB, which were conducted on over 114,000 individuals of European ancestry ^46^.

### Genetic instruments selection

To ensure statistical independence between genetic instruments, we performed a stringent linkage disequilibrium (LD) clumping (window = 10 MB and r^2^ < 0.001 in PLINK 1.9) using the European ancestry data set of the 1000 Genomes Project as a reference panel and selected all the bi-allelic SNPs with minor allele frequency (MAF) > 0.01 using the clump data function in the TwoSample MR R package ^47^. After LD clumping, 208 and 147 SNPs were selected as genetic instruments for puberty timing and adulthood BMI, respectively. Of note, when a SNP instrumenting for the exposure of interest was not available in the outcome summary data, a proxy variant in high-LD with the SNP (window = 1MB and r^2^ > 0.80 in European ancestry) was looked up using the online platform LDlink (https://ldlink.nci.nih.gov/). SNPs would be removed from the sets of genetic instruments if no available proxies can be identified. As no qualified alternative proxy variants were found in each step, the analyses did not use any proxy SNPs. Detailed information on IVs for puberty timing and adulthood BMI is presented in **Table S1-S6**.

## Statistical analysis

### Two-sample MR analysis

As shown in **Fig 1**, a two-sample MR analysis was used as the primary analysis approach to examine the causal effects of genetically predicted puberty timing on 174 human plasma metabolites. The IVW method, which assumes no correlations or interactions between genetic instruments and generates average causal effect estimates across multiple instruments, was used in the primary MR analysis ^48^. The causal effect estimates were interpreted as standardized mean differences in concentrations of metabolites per year delayed onset of puberty. Heterogeneity between causal effect estimates for each SNP was examined by calculating Cochran’s Q statistic, which indicated potentially horizontal pleiotropy.

### MR mediation analysis

To investigate the extent, to which adulthood adiposity might mediate the causal relationship between puberty timing and metabolite levels, we performed an MR mediation analysis (**Fig 1-2**). The indirect causal effects of puberty timing on plasma metabolite levels were estimated using the two-step MR method ^49^. In the first step, we tested the causal effects of adulthood BMI on all plasma metabolite levels. In the second step, we tested the causal effect of puberty timing on adulthood BMI. Then we used the “product of coefficients” approach ^50^ to calculate the indirect causal effects of puberty timing on each blood metabolite via adulthood BMI. The standard error of each indirect causal effect was estimated by using the “multivariate delta method” ^51^.

Compared with conventional univariable two-sample MR, which estimates the total effect of one exposure on the outcome, MVMR includes multiple risk factors as exposures to estimate the direct causal effect on the outcome after adjusting for other risk factors ^52^. We accomplished MVMR analysis to assess the direct causal effects of both puberty timing and adulthood BMI on 174 plasma metabolites. Since only participants involved in the ReproGen consortium were available in the full summary statistics of puberty timing GWAS ^34^, a total of 207 IVs for puberty timing used in the MVMR were directly selected from the summary data following the same LD clumping strategy.

### Sensitivity analysis

To assess the robustness of our results in the two-sample MR analysis, we also performed four other sensitivity analyses. MR-Egger regression provides an unbiased estimate of the causal effect even if all SNPs used as instrumental variables are invalid^53^. The weighted median model requires that at least 50% of the SNPs were valid genetic instruments to provide unbiased estimates of causal effects in MR analyses ^54^. The weighted mode method provides consistent causal effect estimates even if the majority of SNPs were invalid ^55^. The Mendelian Randomization Pleiotropy RESidual Sum and Outlier (MR-PRESSO) method detects outlying genetic variants that might lead to horizontal pleiotropy and further provides causal effect estimates after removing outliers ^56^. Horizontal pleiotropy was tested via the intercept terms in the MR-Egger regression analysis.

As birth weight and childhood obesity were investigated as common causes of puberty timing and some of the adulthood metabolite levels ^57^, to minimize the risk of violating one of the MR core assumptions (i.e., exclusion restriction assumption) ^58^, we excluded SNPs that were genetically significantly associated with birth weight or childhood BMI (*P* < 5 × 10^−8^) from genetic instruments. Based on this, a secondary IVW analysis was performed with the reduced genetic instrument sets.

### Replication analysis in the UK Biobank

The UKB, a population-based cohort, has recruited approximately 500,000 individuals aged 40 to 69 years across the UK. By utilizing the targeted high-throughput nuclear magnetic resonance (NMR) metabolomics platform (Nightingale Health Ltd; biomarker quantification version 2020), 249 metabolic measures (i.e., 165 metabolic measures and 84 derived ratios) were quantified simultaneously ^46^. However, given that only nine of the metabolites (i.e., alanine, glutamine, glycine, histidine, isoleucine, leucine, phenylalanine, tyrosine, and valine) measured in the UKB were identified to coincide with the metabolites included in the primary analysis, we merely estimated the relationship between puberty timing and the nine amino acids to inspect the robustness of our primary analysis results. Firstly, we conducted a two-sample MR analysis to investigate the total effect of puberty timing on each amino acid. Subsequently, we also performed MVMR analysis to obtain the direct impact of puberty timing on amino acids after adjusting for adulthood adiposity. Of 68 SNPs identified as associated with puberty timing, 60 were eventually used in two-sample MR, and the 8 SNPs were removed due to being palindromic with intermediate allele frequencies. A total of 55 SNPs were employed in MVMR analysis after LD clumping.

### Multiple testing correction

Given that a large number of tests were performed, a conservative Bonferroni multiple-test correction was applied. The *P* value threshold for statistical significance becomes *P* < 4.90 × 10^-4^ (0.05/102) after correcting for 102 tests corresponding to the number of principal components explained 95% of the variance of the 174 metabolites in the GWAS by Lotta et al. ^59^, which indicated strong causal evidence for our results. Meanwhile, the conventional *P* value threshold of 0.05 was also reported to avoid ignoring potentially crucial findings after constructing a “suggestive significance threshold” ^60,61^, which offered moderate causal evidence.

All statistical analyses were performed using R version 4.1.2, in which the TwoSampleMR (version 0.5.6) and MRPRESSO (version 1.0) packages were used ^47,56^. The ggpolt2 and circlize R packages were used to visualize statistical analysis results ^62,63^.

## Data Availability

All data produced are available online. The genetic instruments for puberty timing were obtained from the corresponding literature at https://reprogen.org/data_download.html. Data on plasma metabolites GWAS can be found at www.omicscience.org. Full summary data on adulthood BMI were obtained from the MRC Integrative Epidemiology Unit OpenGWAS project (https://gwas.mrcieu.ac.uk/). Full summary data on birth weight and childhood BMI were obtained from the Early Growth Genetics Consortium (egg-consortium.org). The replication analyses were conducted using data from the UKB (https://www.ukbiobank.ac.uk/) and the summary level data were obtained at https://gwas.mrcieu.ac.uk/.

https://gwas.mrcieu.ac.uk/

http://egg-consortium.org/index.html

## Code availability

Scripts are available on GitHub at https://github.com/ZengjunLi/Puberty_metabolites.

## Acknowledgements

This study was supported by grants from the National Natural Science Foundation of China (82373588, 82125032, 81930095 and 81761128035), Xinhua Hospital, Shanghai Jiao Tong University School of Medicine, Shanghai, China (2021YJRC02), Innovative research team of high-level local universities in Shanghai (SHSMU-ZDCX20211100), the Science and Technology Commission of Shanghai Municipality (19410713500 and 2018SHZDZX01), the Shanghai Municipal Commission of Health and Family Planning (GWV-10.1-XK07, 2020CXJQ01, 2018YJRC03) and Sichuan Science and Technology Program (2021JDR0189). The funders of the study had no role in the study design, data collection, data analysis, or data interpretation; writing or reviewing of the manuscript; and decision to submit the manuscript for publication.

## Author information

## Authors and Affiliations

1. Department of Epidemiology and Biostatistics, West China School of Public Health and West China Fourth Hospital, Sichuan University, 610041, China.

Zengjun Li & Cairong Zhu*

2. Population Health Sciences, Bristol Medical School, University of Bristol, Bristol, United Kingdom.

Si Fang

3. Medical Research Council (MRC) Integrative Epidemiology Unit (IEU), University of Bristol, Bristol, United Kingdom.

Si Fang

4. Ministry of Education and Shanghai Key Laboratory of Children’s Environmental Health, Xinhua Hospital, Shanghai Jiao Tong University School of Medicine, Shanghai, 200092, China

Dong Liu, Fei Li* & Jian Zhao*

5. Department of Developmental and Behavioral Pediatric & Child Primary Care, Brain and Behavioral Research Unit of Shanghai Institute for Pediatric Research, Xinhua Hospital, Shanghai Jiao Tong University School of Medicine, Shanghai, China

Fei Li*

6. Department of Maternal and Child Health, School of Public Health, Shanghai Jiao Tong University, 200092, China.

Fei Li* & Jian Zhao*

## Contributions

Cairong Zhu, Fei Li, and Jian Zhao initially conceived the study design. Zengjun Li and Si Fang carried out statistical analyses, and Zengjun Li wrote the first draft of the manuscript with input from Cairong Zhu, Fei Li, and Jian Zhao. Zengjun Li and Dong Liu plotted the figures. All authors contributed to the interpretation of the results and critical revision of the manuscript.

## Corresponding author

Correspondence to Jian Zhao, Cairong Zhu, and Fei Li.

## Ethical approval

This study used publicly available GWAS summary data and was not subject to institutional review board approval.

## Competing interests

The authors declare no competing interests.

## Supplementary information

Table S1-S21.

## Notes

### Competing Interest Statement

The authors have declared no competing interest.

### Author Declarations

The study used (or will use) ONLY openly available human data. The genetic instruments for puberty timing were obtained from the corresponding literature at https://reprogen.org/data_download.html. Data on plasma metabolites GWAS can be found at www.omicscience.org. Full summary data on adulthood BMI were obtained from the MRC Integrative Epidemiology Unit OpenGWAS project (https://gwas.mrcieu.ac.uk/). Full summary data on birth weight and childhood BMI were obtained from the Early Growth Genetics Consortium (egg-consortium.org). The replication analyses were conducted using data from the UKB (https://www.ukbiobank.ac.uk/) and the summary level data were obtained at https://gwas.mrcieu.ac.uk/.

